# Comparing Heart Rate Variability in Canadian Armed Forces Patients to Control Participants without Chronic Pain/Mental Health Issues

**DOI:** 10.1101/2022.05.15.22275102

**Authors:** Latifah Kamal, Amir Minerbi, Tali Sahar, Keri J. Heilman, LCol Markus Besemann, Vidya Sreenivasan, Salena Aggerwal, Gaurav Gupta

## Abstract

**Background:** The autonomic nervous system is subserved by the sympathetic and parasympathetic which regulate vital involuntary physiological functions like heart rate. Parasympathetic activity can be measured from the high-frequency component of heart rate variability (HRV), measured via the amplitude of RSA, as a possible predictor for mental health and chronic pain disorders. Therefore, investigators looked to correlate HRV with chronic pain when compared healthy controls.

**Methods:** As part of a larger ongoing study, patients complete pre-defined questionnaires on their pain condition, potential risk factors, and function. For patients and controls investigators collected performance and cardiac measures (RSA, LF-HRV, heart period) while at rest, walking and lifting tests. This analysis focused on differences in heart rate variability measures between 100 patients and 48 controls.

**Results:** Preliminary analysis revealed demographic and anthropometric variables varied significantly between groups. When comparing HRV measures, respiratory sinus arrhythmia (RSA) during lying and sitting were significantly decreased in patients compared to controls while heart period lying and walking were significantly increased in patients. Correlation analysis revealed significant positive correlation between RSA during lying and sitting when looking at age, gender, and weight. Heart periods during lying and walking were negatively correlated with gender and weight.

**Discussion:** To our knowledge it is the first study to look at chronic pain and HRV in the Canadian Forces, while also collecting data on patient reported outcomes, and during various resting and activities. Many potential limitations exist for this study including challenges with respect to controlling for known confounders of heart variability.

**Conclusion:** By establishing heart rate variability as a correlate of chronic pain, the outcome of this project could potentially improve quality of care for patients with these conditions. Further work controlling for confounders and relating HRV to pain severity, subtypes, patient reported outcomes and functional abilities will be required to determine the exact value for clinical decision making.

## Background

Up to 20% of Canadians suffer from chronic pain with much higher estimated prevalence rates in older adults and those in long term care [1,2,3]. Chronic pain is often unrecognized and/or undertreated which, as a result, has significant impacts on functional abilities, quality of life, societal participation and health care utilization [4,5,6,7,8]. Even when compared with other diseases, quality of life among patients suffering from chronic pain is poorer [4], with associated productivity costs and health expenditures being up to $60 billion in Canada, and $635 billion in the United States per year [9,10].

While epidemiologic information regarding Canadian civilians and chronic pain exists [8], information regarding the Canadian Armed Forces (CAF) is limited. In CAF veterans, 41% of the population experienced constant pain and 23% experienced intermittent chronic pain [11]. When compared to the Canadian civilian population, the prevalence of activity reduction for CAF veterans was remarkably higher (49% versus 21% among civilians), and a greater percentage of individuals in the veteran population required assistance with at least one activity of daily living (17% versus 5% among civilians) [12]. Disability odds were higher for veterans with chronic pain (10.9, p<.05), than mental health conditions (2.7 p<0.05), but unsurprisingly there was a combined impact of physical and mental health issues [12]. In the United States, certain veteran populations have a chronic pain prevalence of about 47%, with moderate-to-severe pain in 28% [13]. Therefore, mental health and musculoskeletal/chronic pain issues are a leading cause of functional limitations, decreased quality of life, and economic loss in patients leaving military service [14].

The current assessment of functional status in patients with chronic pain relies primarily on patient reported outcomes in the absence of objective standard measures of physical function. Since patients with chronic pain have a complex interplay of non-reversible health issues, medications and psychosocial issues, and having a clear and objective understanding of pain is of paramount importance in pain reduction targets and activity optimization. Therefore, developing a physiologic correlation of physical and emotional capacity with respect to pain and function would be beneficial.

Both sympathetic and parasympathetic nervous systems are involved in regulating pain states and may be disturbed in chronic pain. Autonomic dysfunction, including an increase in resting heart rate and reduced heart variability can occur in various chronic pain states [15, 16, 17, 18, 19, 20].

Heart rate variability (HRV) is considered a sensitive predictor of the capacity to regulate emotional responses to threatening intrinsic and extrinsic stressors, and low HRV is associated with poor long-term health outcomes in various health states including cardiovascular disease and mood disorders. High heterogeneity aside, pooled results from one meta-analyses reflected a consistent, moderate-to-large effect of decreased high-frequency HRV in patients with chronic pain, implicating a decrease in parasympathetic activation [21,22]. Researchers have also developed several vagal tone (VT) indices and formulated the polyvagal theory that provides a fruitful ground for the research, understanding, and treatment of different chronic medical and mental health conditions [23, 24].

There are various measures of heart rate variability and this analysis focused on range reflecting various aspects of autonomic control. The time distance between successive QRS complexes (measured using R-R intervals) is the heart period, which reflects all contributions to overall HRV. Respiratory sinus arrhythmia (RSA), the primary component of high frequency HRV (HF-HRV), is a potential measure of parasympathetic control between heart beats coordinated with respiration [25]. RSA forms a portion of brainstem nuclei output measurements along with the fluctuations in heart rate associated with spontaneous breathing [35]. The amplitude of RSA values obtained show a relationship with the cardiac vagal tone which has an inhibitory effect on the sinoatrial node of the heart [35]. Low-frequency HRV was also analyzed in the current study as a potential measure of blood pressure/baroreceptor activity [38]. This study therefore aims to determine if the heart rate variability measurements can help to distinguish patients with chronic pain from healthy controls. Investigators hypothesize that patients with chronic pain will have different HRV measurements than the controls.

## Methods

Between October 2021 and February 2022, 100 actively serving members of the Canadian Armed Forces (CAF) with chronic pain and 50 pain-free controls from the Canadian Forces Health Services Centre - Ottawa (CFO) and community were recruited for participation. Approval from the Defence Research Ethics Board, Canadian Forces Surgeon General, and Commanding Office at CFO was obtained prior to commencement.

Inclusion criteria were male and female between 18 and 70 years old, with patients being seen within the CFO and diagnosed with one or more chronic pain conditions. Exclusion criteria were participants or guardians unable to give informed consent, adults with uncontrolled mental health issues, pacemaker or active cardiac Issues, and specific medication use in controls (i.e. Serotonin Reuptake Inhibitors/Beta Blocker/Anticholinergic, migraine, cold medication, antihistamine, and cannabis use). Control participants between 12-18 years old were able to participate with parental consent.

This study on the differences between patients with chronic pain and pain-free controls was part of larger analysis looking at patient reported outcomes, functional and physiological measures in patients with chronic pain when compared to controls. For this specific investigation, we included information on whether the participants had chronic pain, their age, gender, height, weight, medications taken, and recent exercise, food consumption, smoking habits, cannabis and caffeine intake as possible confounders.

The entire data set includes demographics (i.e. Age, gender, rank, working status, medical category), height and weight, current medications (i.e. Antipsychotics, antihypertensives, antidepressants, oral contraceptive pill, stimulants and cannabinoids), activity and food intake (i.e. Alcohol, caffeine, energy drinks, food, intense exercise, tobacco, cannabis). Patient reported outcomes taken at baseline include body pain diagram, Pain, Enjoyment and General Activity Scale (PEG), Pain Disability Index (PDI), Hospital Anxiety and Depression Scale (HADS), Adapted Deployment Risk & Resilience Inventory 2 Section J: Unit Support Exercise Questionnaire, Brief Trauma Questionnaire (BTQ) and Litigation Status.

The physical activity measures taken include the 2 minute walk test (2MWT) and Elevation & Movement Lift test (EMLi). Other data included cardiac measures (heart period, RSA and LF-HRV) and Borg Perceived Exertion Scale. The positional and active tasks performed by participants were as follows: Supine (3 minutes), Sit (3 minutes), Stand (3 minutes), EMLI (1 minute), 2MWT (2 minutes). This information will allow for follow-up analysis and to further control for known confounders [26, 27, 28]. Investigators used the Firstbeat Bodyguard 2 (Firstbeat Finland) for acquisition of electrocardiogram data (ECG) with data sampled at 1000 Hz [29].

The 2MWT was conducted by asking participants to walk at moderate intensity (brisk walk) for the duration of the activity (2 minutes). The distance walked was recorded in meters. The EMLi test consists of pushing a 2kg weighted ball from the chest level forward, and then raising the ball overhead, before returning it to the starting position (Figure 1). The number of repetitions done within 1 minute at moderate intensity was recorded.

**Figure 1.** Demonstration of the EMLi test where a 2kg weight ball is pushed from the chest level forward (A to B), then raised overhead (C to D), before returning it to the starting position (A).

For Figurer 1 pls see https://www.healthaya.com/publications

## Results

Upon recruitment of the sample size total 150 participants, data on 100 chronic pain patients and 48 controls were able to be analyzed. After collection, 2 controls were excluded because of high intensity exercise and alcohol use in the last 24 hours. No patients had associated cardiac disease or pacemaker, 27% were taking SSRI/anticholinergics/ beta blockers and 14% had taken migraine, cold medications and/or antihistamines in past 48 hours. Additionally 4% were taking antipsychotics, 19% antihypertensives, 13% cannabinoids, 2% birth control, and 6% stimulants. Finally 20% of patients used alcohol in the previous 24 hours, 42% used caffeine products in the previous 2 hours, 6% percent used energy drinks in the previous 48 hours, 44% ate food in the previous 2 hours, 12% did intense exercise in the previous 24 hours, 8% used tobacco products/smoked in the previous 2 hours and 1 person used plant based cannabis in the previous 2 hours.

There were no adverse events noted. Age, gender distribution, and anthropometri variables varied significantly between groups. When comparing HRV measures, RSA1-Lyin and RSA2-Sitting were significantly decreased in patients as compared to controls, while heart period 1-Lying and 5-Walking were significantl increased in patients. There were no correlations noted when comparing low frequency heart rate variability. Correlation analysis revealed significant positive correlation between RSA1-Lying and RSA2-Standing, with age, gender and weight demographics. Heart periods 1-Lying and 5-Walking were negativel correlated with gender and weight. See Table 1 and Figure 2 for comparison of results.

**Table 1:** Results Comparing Demographics and Various Heart Rate Variability Measures Between Patients with Chronic Pain and Controls.

| Variable | Mean Patients | Std Dev Patients | Mean Controls | Std Dev Controls | P Val | FDR |
| --- | --- | --- | --- | --- | --- | --- |
| Age | 43.81 | 7.76 | 36.79 | 16.19 | 0.000 | 0.001* |
| Gender (F:M) | 32:68 | --- | 30:18 | --- | 0.0004 (chi<sup>2</sup>) | 0.000* |
| Height (in) | 69.23 | 9.82 | 65.49 | 3.83 | 0.000 | 0.000* |
| Weight (lbs) | 190.00 | 38.65 | 150.27 | 35.20 | 0.000 | 0.000* |
| RSA 1 – Lying | 5.70 | 1.45 | 6.53 | 1.81 | 0.003 | 0.017* |
| RSA 2 – Sitting | 5.34 | 1.56 | 6.09 | 1.51 | 0.007 | 0.026* |
| RSA 3 – Standing | 4.49 | 1.19 | 4.86 | 1.55 | 0.112 | 0.280 |
| RSA 4 – EMLi | 4.36 | 2.52 | 3.90 | 2.31 | 0.300 | 0.421 |
| RSA 5 – Walking | 4.54 | 3.43 | 4.68 | 3.66 | 0.813 | 0.813 |
| Heart Period 1 - Lying | 964.89 | 665.56 | 935.76 | 155.83 | 0.293 | 0.421 |
| Heart Period 2 - Sitting | 876.53 | 355.94 | 885.95 | 154.54 | 0.148 | 0.318 |
| Heart Period 3 - Standing | 749.68 | 111.54 | 772.52 | 153.78 | 0.309 | 0.421 |
| Heart Period 4 – EMLi | 660.68 | 202.61 | 558.55 | 86.50 | 0.001 | 0.010* |
| Heart Period 5 - Walking | 574.97 | 113.63 | 505.88 | 92.33 | 0.000 | 0.006* |

Note: \* statistical significance. Means were collected using Pearson's Chi-squared test.

**Figure 2:**
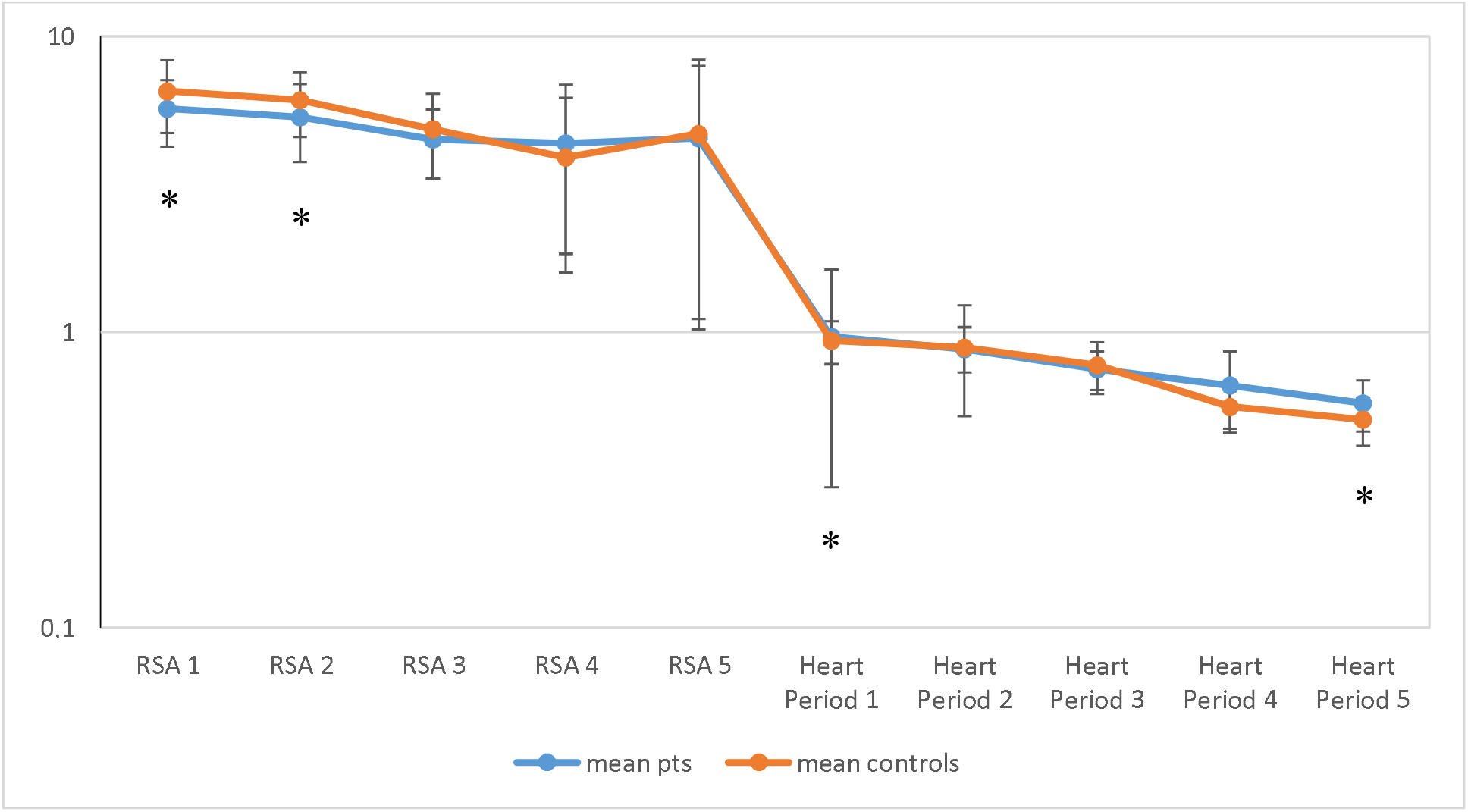
Results Comparing Various Heart Rate Variability Measures Between Patients with Chronic Pain and Controls. Note: * statistical significance. Values for Heart Periods 1-5 seen in Table 1 were divided by 1000 for scaling in the figure above.

Table 2 depicts RSA confounder criteria met by participants in the patient group. Notably, 55% of the participants in this group had at least 1 confounder criteria, and 22% had more than one confounder criteria. These results imply that over half of the chronic pain patients studied are additionally dealing with factors that may impair vagal modulation.

**Table 2:** RSA Confounder Criteria Met By Patient Group.

| No Confounder (0) | 1 | 2 | 3 | 4 | 5 | Any Confounder |
| --- | --- | --- | --- | --- | --- | --- |
| 45 | 33 | 14 | 5 | 2 | 1 | 55 |

## Discussion

This study is unique for many reasons. To our knowledge, it is the first to look at chronic pain and HRV in a Canadian military setting, while also collecting data during various rest positions, activities, and on patient reported outcomes (to be subsequently analyzed). While statistical significance for specific RSA and HP measures were seen for specific positions/activities, the clinical significance, effect size and relative impact of confounders requires further work [30].

Previous research has also shown that reductions in HRV are correlated with perceptions of activity impairment [31]. The directionality of this relationship is yet unknown, specifically whether chronic pain and autonomic dysregulation are correlates of another mechanism, or related by cause and/or effect [22]. However, some authors argue that irrespective of the mechanism, “both implicate a greater risk of mortality and disease beyond the direct impact of pain, warranting systematic evaluation of the evidence, before recommending targeted management” [22].

The utility of low frequency – heart rate variability is still up for debate. While comparing across units of measurement may not provide validity, logarithmically transformed HRV could be useful, and represent either sympathetic activation, and/or baroreflex related cardiac response [32, 33, 34, 35, 36, 37, 38]. Alternately, there appears to be moderate-to-large effect size for HF-HRV reduction compared with healthy controls. While the studies focused largely on patients with fibromyalgia, this suggests a decrease in parasympathetic activation, which is already associated with poor health outcomes [39, 40, 41, 42, 43, 44, 45, 46]. Finally, RSA, or vagally mediated HRV – regarded as the central regulation of peripheral function, is hypothesized to be impaired in chronic pain states, corresponding to descending spinal inhibitory control and central sensitization [47, 48, 49, 50, 51, 52].

Many potential limitations exist for this study. Confounders for HRV general are challenging to control. Sinoatrial node influence on HRV can be modified by many factors such as age, gender, anthropomorphic, thermoregulation, the sleep-wake cycle, hormones, stress, physical activity, substance use and medications.

Controlling for confounders, especially medication use amongst chronic pain patients, served to be a significant challenge and was therefore not strictly controlled. Controlling for extrinsic factors such as noise was much easier however certain instances of acute loud noises did occur beyond the investigators’ control. Sudden loud noises are known to cause increases in heart rate [67]. One significant issue is not controlling for the type, location and/or number of sites for patients with chronic pain. Though we have acquired this information from the participants in this study for further review, it appears that chronic pain, irrespective of its etiology and phenotypes, share many similar mechanisms [53].

Age and gender were shown to be correlated with HRV in this study. This is consistent with previous literature that shows HRV declining by as much as 15% for each decade, and pre-menopausal gender differences exist between males and females [54, 55, 56, 57, 58]. Unlike previous literature and the control group however, patients in this study were more likely to be older and male due to the population in which the study was conducted.

Cardiac disease, psychiatric conditions, certain medications (e.g. Tricyclic antidepressants, selective serotonin reuptake inhibitors, calcium channel, or beta-blockers) and lifestyle choices (e.g. Caffeine and nicotine) are known to impact heart rate variability [59, 60, 61]. While these were documented for patients and strictly enforced for controls, after screening close to 300 patients, we believe controlling for these factors can only occur with extremely large data sets and higher level statistical analysis/machine learning models in future work. Further analysis will look at multivariate analysis to determine the relative impact of these factors, and their correlation to pain and physical function.

In the future, using patients as their own controls and looking at HRV over time and/or in response to treatment might be the only way to provide robust and valuable clinical insights. If a relationship can be established, it can potentially help identify patients at risk for developing chronic pain, guide treatment decisions along with testing new therapies and technologies in a personalized way. Vagal efficiency will be calculated by the slope between the dynamic and synchronous shifts in RSA and heart period, which provides a measure of heart rate regulation by vagal pathways [36].

## Conclusion

Patients with chronic pain have specific cardiac measures (RSA and heart period) that are different than in controls for specific positions/activities. The preliminary outcomes are confounded by the impacts of demographic and clinical factors that require further analysis and larger datasets for precise results before concluding the presence and/or degree of autonomic dysfunction in patients with chronic pain. By establishing heart rate variability as a correlate of chronic pain, the outcome of this project could potentially improve the quality of care for patients with these conditions. Further work controlling for confounders, and relating HRV to pain severity, subtypes, patient reported outcomes, and functional abilities will be required to determine the exact value of HRV measurements for clinical decision-making.

## Disclaimer

The views expressed herein are those of the authors and may not reflect those of the Canadian Armed Forces, Canadian Forces Health Services, the Department of National Defence or the Canadian Government.

## Data Availability

Data not available

